# Short term atrial fibrillation in different stroke entities

**DOI:** 10.1101/2023.09.28.23296319

**Authors:** Priyanka Boettger, Till Kracht, Nils Schulz, Kai Ott, Philipp Klemm, Uwe Lange, Ulf Müller-Ladner, Andreas Rolf, Henning Lemm, Michael Buerke

## Abstract

**Background:** Stroke including ESUS is one of the most common causes of disability and death in the world. Atrial fibrillation (AF) of more than 30 sec is considered a reason for cardioembolic stroke. Nevertheless, shorter periods of AF might also be a possible risk factor.

**Methods:** We determined periods of AF within a range of 0-14 s, 15-30 s and > 30 s in patients with different stroke entities. We analysed all ischemic strokes and transient ischemic attacks at the Stroke Units/Emergency Room in Siegen within 6 months and classified them etiologically. All stroke patients were prospectively ECG monitored over a minimum of 24h and characterized for the different AF periods at the stroke unit. The stroke entities were then compared.

**RESULTS:** A number of 714 patients over 6 months were hospitalized suffering a stroke. Among them 163 (30.8%) had a cryptogenic Stroke, including 98 (19%) ESUS patients. Furthermore, 185 (26%) TIA, 209 (39%) cardioembolic, 110 (21%) atherosclerotic, 40 (8%) lacunar and 7 (1%) other specific strokes were registered. Manifest atrial fibrillation (>30s) showed a prevalence of 23% within our stroke population. Whereas 15% had an AF episode of 15-29 s and 16% presented AF episode of 0-14 s. Among cardioembolic infarcts, 45% had manifest atrial fibrillation (>30 s), 20% had an atrial fibrillation episode of 15-29 s, and 22% showed an atrial fibrillation episode of 0-14 s. Taken together, more than 90% of cardioembolic infarcts show an episode of atrial fibrillation of any duration. An AF episode of 15-29 s or 0-14 s was also found in almost 35% of ESUS patients: similar to cryptogenic infarctions. In 22% of the TIA patients manifest atrial fibrillation had been detected. 7% of TIA patients showed one or more episodes of AF of 15-29 s and 10% an episode of 0-14 s. Among the atherosclerotic infarctions, the proportion of AF >30 s is profoundly lower at 12%, however the percentage of AF for 15-29 s was 14. The smallest ratio of atrial fibrillation of any duration was formed by lacunar insults with a total of nearly 25%, of which only 13% had manifest AF.

**Conclusion:** Non-valvular atrial fibrillation presents the most common cause of cardiac cerebral embolism. In our study, AF showed the largest ratio among cardioembolic strokes with 48%. Overall, AF of any duration occurred in nearly 90% of this group. Since a manifest AF is an exclusion criterion for ESUS, conversely, none of the ESUS patients suffered from manifest AF. However, it is striking that 15% of ESUS patients had an AF episode of 15-29 s and 18% had an AF episode of 0-15 s. Similar results were observed in cryptogenic strokes. Possibly, the definition of AF has to be adjusted, since the stroke subgroups with short term AF did not receive proper anticoagulation treatment. However, at least advanced rhythm monitoring should be provided. This should be investigated in future prospective studies.

## INTRODUCTION

Atrial fibrillation (AF) is the most common cardiac arrythmia and a major cause of stroke, heart failure, sudden death, and cardiovascular morbidity. AF increases risk of thromboembolic stroke via stasis in the left atrium and subsequent embolization to the brain. Atrial fibrillation may be asymptomatic and consequently subclinical. ^1^ Epidemiologic studies indicate that many patients with atrial fibrillation on screening electrocardiograms had not received a diagnosis of atrial fibrillation previously. ^2^ About 15-23% of all strokes are attributable to documented atrial fibrillation, and 50 to 60% to documented cerebrovascular disease. ^3, 4^ In about 25% of patients who have suffered from ischemic strokes, no etiologic factor is identified (Jabaudon2004). Subclinical atrial fibrillation is often suspected to be the cause of stroke in these patients. ^5^ The minimum duration of an ECG tracing of AF required to establish the diagnosis of clinical AF is at least 30 seconds, or entire 12-lead ECG. ^6, 7^ In patients with acute ischemic stroke, it is essential that clinicians undertake careful investigation to detect subclinical AF. In these patients, up to 23.7% are eventually found to have underlying AF. The initial purpose of the TOAST classification system was to categorize stroke patients more precisely in purpose of investigating any potential efficacy of LMWH anticoagulant for treatment of various types of ischemic strokes. ^8^ This system was primarily based on clinical features plus any information from neuroimaging, echocardiography, neurosonography or cerebral angiography. The TOAST classification system has been used extensively for other purposes, such as identifying new genetic markers and risk factors. ^9^ The TOAST system was composed of five major subtypes: large artery atherosclerosis, cardioembolism, small artery occlusion, stroke of other determined cause (SOC), and stroke of undetermined cause. The toast classification leaves a subgroup of 25-30%, ^10^ ^11^ whose cerebral insults are classified as cryptogenic. The so-called cryptogenic strokes can either be caused by missing diagnostic clarification, several competing mechanisms or can actually remain cryptogenic, i.e. of unknown etiology. The TOAST criteria provide no minimum diagnostic standard for the classification of stroke, consequently the classification was questioned for the first time in 2014 and a new diagnostic-oriented concept called ESUS was introduced by the "Cryptogenic Stroke/ESUS International Working Group". The underlying idea was a classification providing a more specific distinction between stroke entities after excluding precisely defined stroke causes, thus giving the former cryptogenic stroke a new face called ESUS. ^10^ According to current standards, the etiological assignment is based on a diagnostic minimum standard and consists of the following examinations: cerebral imaging, electrocardiogram, echocardiography, and imaging of the extra- and intracranial vessels. For the definitive diagnosis of an embolic stroke of unclear etiology according to ESUS criteria, the following causes of the stroke have to be excluded: macroangiopathic genesis, cardiac embolism, atrial myxoma or other cardiac tumors, a high grade mitral valve stenosis, a myocardial infarction less than 4 weeks ago, a left ventricular ejection fraction <35%, valvular vegetations or an endocarditis, a microangiopathic genesis, and other defined causes (vasculitis with cerebral involvement or a dissection of the arteries supplying the ischemic cerebral area, migrainous infarctions, arteritis, prothrombotic coagulation disorders, angiological relevant storage diseases such as Fabry’s disease and the reversible cerebral vasoconstriction syndrome (RCVS)). 7-25% of all ischemic strokes following completion of diagnostics in the acute phase are declared as ESUS. ^12^ ^13^ With a stroke recurrence rate of 3-6% per year, ^10^ the suspicion arises that a relevant part of the strokes initially classified as ESUS is apparently caused be cardiac embolism.

## CARDIAC DIAGNOSTICS AFTER ACUTE ISCHEMIC STROKE

In patients with ischemic stroke, 12-lead resting ECG, continuous ECG monitoring on the stroke unit or long-term ECG registration is recommended to detect relevant cardiac arrhythmia or an acute or past myocardial infarction. The current guideline of the European Society of Cardiology (ESC) also recommends ECG monitoring after an ischemic stroke for a minimum of 72 hours. ^14^ By consensus an atrial fibrillation episode of >30 s is required for the diagnosis of AF. Whether or not periods of <30s lead to manifest or paroxysmal AF in the future or might already form a risk factor for future embolisms is unknown.

Additionally, an ultrasound examination of the heart (echocardiography) should be performed considering the principle recommended above. The non-invasive transthoracic Echocardiography (TTE) and semi-invasive transesophageal echocardiography (TEE) are mandatory tools mentioned in the ESC guidelines for stroke. Today, cardiac MRI has emerged as the examination of choice for detecting an acute myocarditis, cardiomyopathy or heart failure with a calculation of the left ventricular ejection fraction, as well as an identification of coronary heart disease.

## OBJECTIVE AND QUESTION

The present study aims to contribute to the clinical relevance of AF detection in different stroke entities. Currently, the definition of AF requires an AF period of 30 s. However, it is unclear if short term AF might form another risk factor for developing future manifest or paroxysmal AF or cause cerebral embolism itself. Prior studies have proven the stroke prevention of DOAC therapy in AF patients, but prior trials have failed to show that DOAC therapy prevents stroke recurrences in ESUS. ^15^ ^16^ We propose that ESUS encompasses multiple underlying etiologies requiring different types of therapies-among them a subgroup suffering from short term AF. In this study we want to define embolic risk factors and detect short term AF by analyzing the ECG monitoring in stroke patients of all entities.

## METHODS

### Study Population and Definitions

Over 6 month all patients at Siegen hospital which were referred to the stroke center due to a suspected ischemic stroke or who were admitted as the emergency room were examined. The study was conducted in accordance with the Declaration of Helsinki and approved by the Ethics Committee of the Medical Association of Westphalia-Lippe (file number 2015-091-fS). All data (demographics, Comorbidities, clinical and diagnostic values) were recorded prospectively. Patients who were admitted to ER, ICU or the Stroke Unit with the diagnosis of an acute ischemic stroke or a transient ischemic attack and met the inclusion criteria were prospectively included in the study after a clinical examination and followed up until discharge.

**Table 1:** Inclusion and exclusion criteria.

| Inclusion criteria: | Exclusion criteria: |
| --- | --- |
| • Age ≥ 18 years | • Hemorrhagic stroke |
| • First or recurrent event of TIA or acute ischemic stroke | • Hospitalization <24 h |

The neurological deficit of the patients was recorded on admission to the stroke unit/ICU and quantified by the National Institutes of Health Stroke Severity Scale (NIHSS). ^17^ This scale is intended to indicate the severity of neurological deficits. Patients with stroke were surveyed in order to document the clinical course and any drug or interventional treatments. 13 categories were considered, which are among other things, vigilance, orientation, motor skills, sensitivity, visual deficits, language formation and understanding.

NIHSS scores were reassessed every six hours to quickly detect clinical changes in neurological status. The NIHSS has high reliability and validity as well when surveyed by non-neurologists ^18^ ^19^ and is used in clinical stroke diagnostics, therapy, but also in clinical trials. For our study, the score on admission and at discharge was crucial for the classification. It was of interest whether the severity of the clinical manifestation appeared as a correlate of the size of cerebrovascular damage and whether differences are recognized in relation to the neurological deficits and their possible regression in relation to the stroke etiology.

All patients had a 12-lead ECG on admission, 24-hour Holter ECGs during hospitalization and/or ECG monitoring in the ICU or stroke unit.

### Risk factors

There is a number of risk factors that can lead to cardiovascular injury. In addition, cardiovascular diseases and embolic risk factors were recorded in order to assess the risk of developing the various strokes.

**Table 2a:** Cardiovascular risk factors.

| **Risk Factors** | **Parameters** |
| --- | --- |
| Nicotine abuse | > 5 pack years |
| Hypertension | > 135/90mmHg |
| Dyslipidemia | HDL<40 mg/dL, LDL>160 mg/dL, triglycerides>200 mg/dL, or total cholesterol > 240 mg/dL. |
| Overweight | BMI>25kg/m<sup>2</sup> |
| Diabetes mellitus Type I or II | HbA<sub>1c</sub> level of > 6.5% or fasting glucose of 100-126 mg/dL |

**Table 2b:** Embolic risk factors.

| **Risk factors** | **Parameters** |
| --- | --- |
| Previous stroke or TIA | Documented TIA/stroke or on imaging detected old apoplexy |
| Coronary artery disease | Coronary artery stenosis >50%, previous STEMI, NSTEMI or PCI |
| Atrial fibrillation | 1 episode >30s |
| Heart valve prosthesis | Biological or technical heart valve replacement |
| Severe heart failure | EF<35% |

Similar CHA_2_DS_2_VASC score was determined in all stroke groups to characterize further risk of the stroke patients.

## STATISTICS

Based on tabular listings in digital form using Excel® and graphically representations, a descriptive summary of the data could be created. Descriptive statistics were performed for different variables.

## RESULTS

### Clinical characteristics of the patients

From March to August 2015, 714 (+57) patients with suspected stroke were admitted to the Hospital. Among these 57 were strokes of hemorrhagic origin; they were not included in the study. The total number of patients included in the study was n=714. Among them, n= 185 patients had no ischemic area in cerebral imaging, no longer showed any neurological deficit within 24 hours, and were declared as TIA. A stroke of cardioembolic origin was reported by n=209 and n=110 stroke patients suffered from a macroangiopathic stroke. The smallest group was formed by the lacunar strokes (n=40) and other specific causes (n=7). In n=163 patients the stroke cause remained unclear. Within the subgroup of cryptogenic strokes, n=98 patients fulfilled the diagnostic standard for the declaration being an ESUS.

### Risk factors in the overall population

Hypertension being the most common risk factor, occurred in 74% of the total population (strokes and TIA), followed by obesity, which affected almost 50% of the population. The third most frequent factor was hypercholesterolemia, which appeared in almost 40% of the strokes. Other risk factors were CHD, stroke or TIA in the history, diabetes mellitus and nicotine abuse, which predominated in 25-30%. Artificial heart valves and severe heart failure with a left ventricular ejection fraction below 35% was only described in <10% of the stroke patients. (Table 3)

**Table 3.**
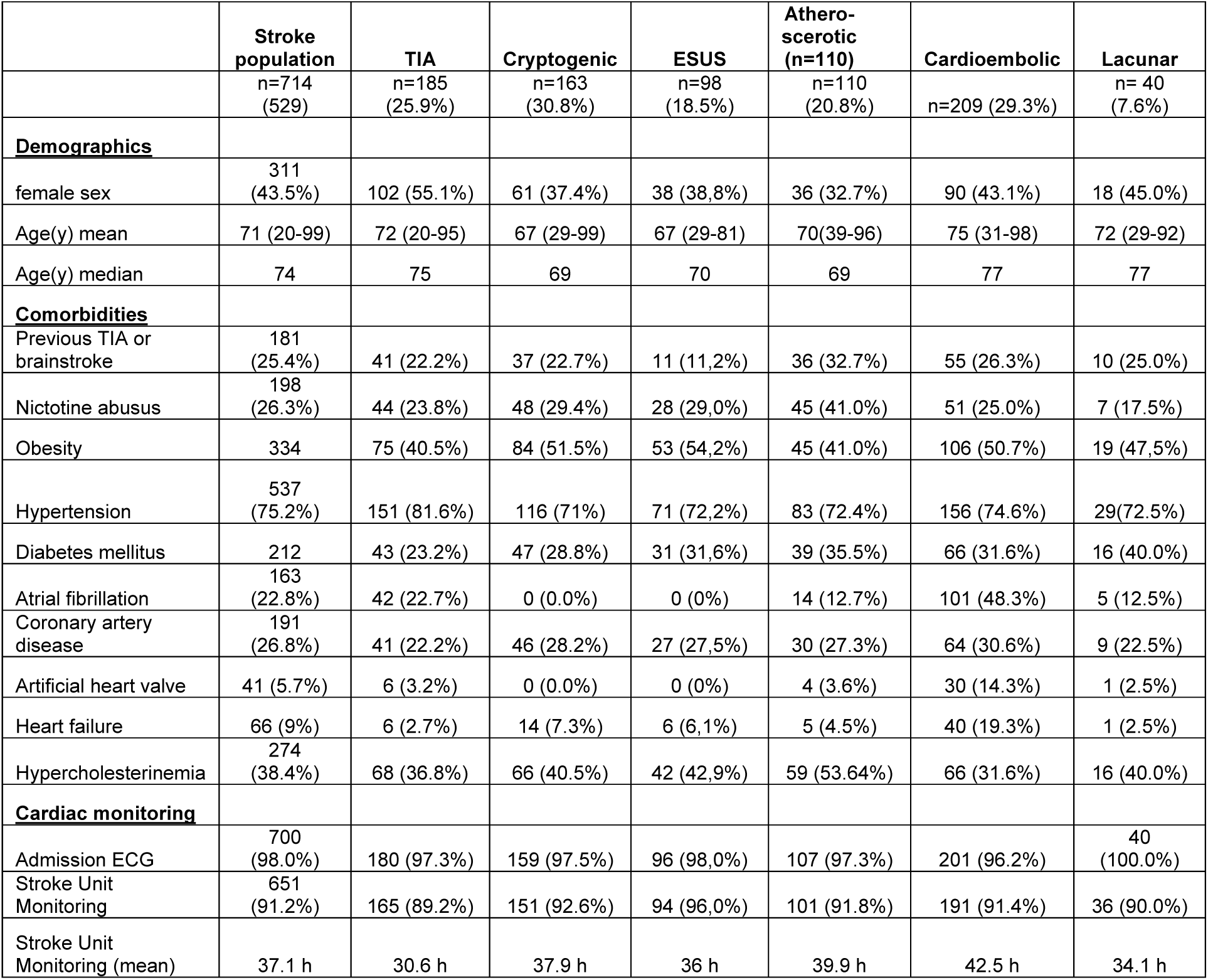
Baseline characteristics and risk factors of patients with ischemic stroke including cryptogenic stroke and ESUS. (Absolute and relative)

Considering the cardioembolic risk factors, CHD with 27% was the most common. 25% suffered from a recurrent stroke or TIA. Half of the patients suffering from cardioembolic infarctions and one fifth of the total stroke patients were diagnosed with the major embolic risk factor AF prior to hospitalization. Among ESUS and cryptogenic stroke, these two risk factors were not recorded, since these, like an artificial heart valve prosthesis, are a part of the ESUS exclusion criteria. Among these stroke entities ischemic heart disease was the leading embolic risk factor.

Hypertension leads by far among the cardiovascular risk factors. Over 70% of patients in each entity suffer from hypertension. More than two-third of these patients were on no or insufficient antihypertensive treatment before admission. The second most common cardiovascular risk factor was obesity. It is most common among ESUS patients at 55%. Hypercholesterolemia was the third most common risk factor in the overall population and was most prevalent in atherosclerotic infarcts at 55%, secondly in ESUS patients at 42%. 26% of total population were smokers. Among the atherosclerotic infarctions the highest relative number of smokers were over 40%. Patients with lacunar infarctions have the smallest proportion of smokers. On the other hand, they have the largest relative number of patients suffering from diabetes at 40%. The second most common manifestation of diabetes mellitus occurred in the stroke patients suffering from macroangiopathic infarctions. The lowest relative number for diabetes mellitus is presented in the TIAs.

### Age groups in each stroke entity

Figure 1 shows the distribution of the proportion of each stroke entity in four age groups. Among the youngest age group (<65 years) over 30% were cryptogenic/ESUS and 25% suffered from a TIA. The distribution of stroke entities in middle-aged patients (66-74 years) was equal. Any stroke entity (other than lacunar infarction) reaches 20-25%. Among the 75- to 84-year-olds, the largest group was formed by cardioembolic strokes with almost 40%, followed by TIA with almost 25%. Cryptogenic strokes and atherosclerotic infarctions account for the smallest proportion. Cardioembolic stroke was also most prevalent among patients >85 years old reaching over 45%, followed by the TIAs with 33%. (Figure 1)

**Figure 1:**
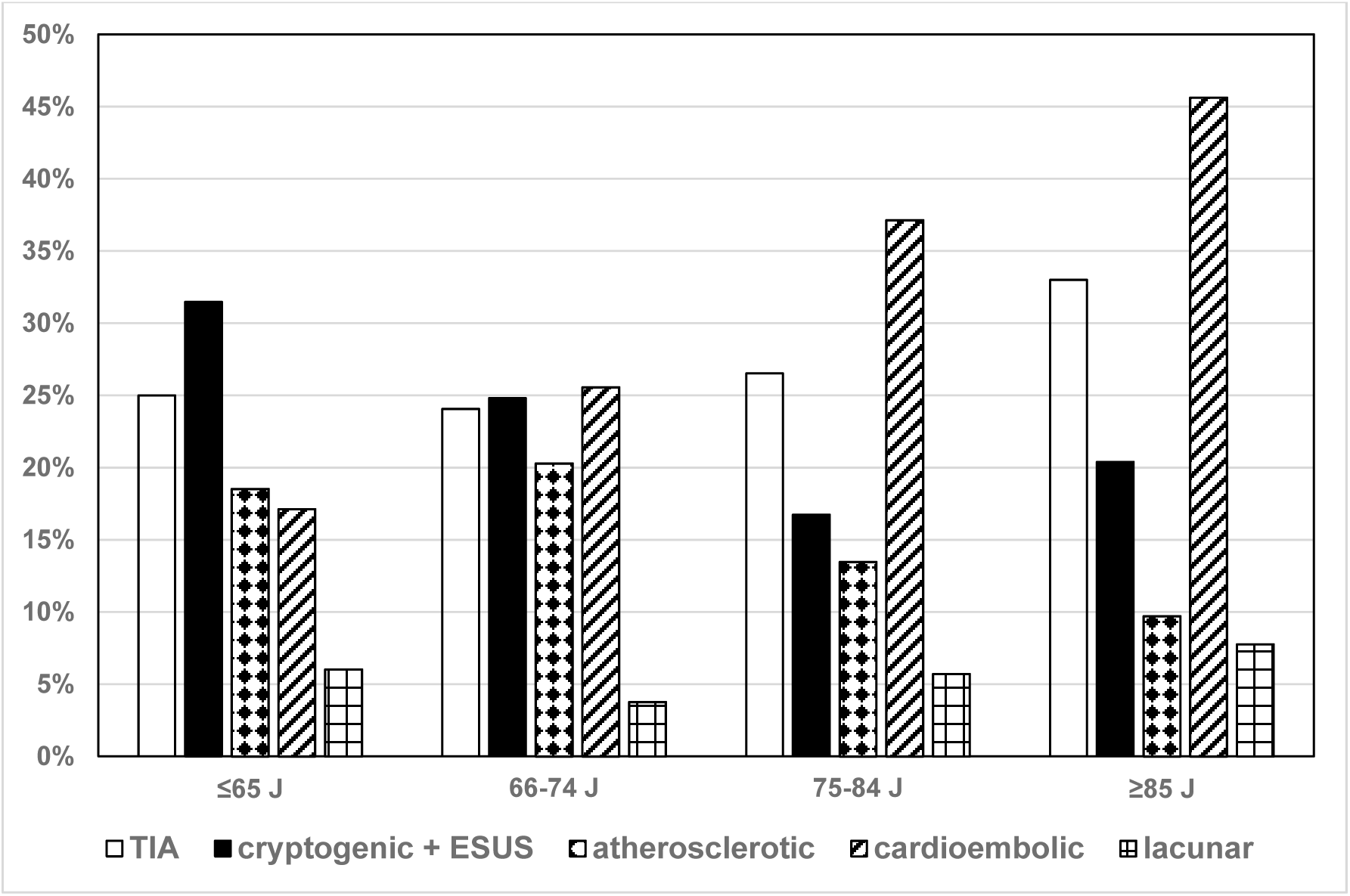
Distribution of stroke entities within each age group

### Stroke Severity

The severity of stroke was recorded using the NIHS score. This was clinically collected within 24 hours after admission and before discharge. The following results were found: In general, there was an overall decrease of the neurological deficits at discharge. The NIHSS on onset of neurological symptoms with an average of 7 points was importantly higher than at discharge into a suitable rehabilitation measure (Average NIHSS of 3 points). On average, the NIHSS is halved from admission to discharge. Patients suffering from cardioembolic infarction (mean NIHSS on admission 11) have the largest neurological deficits, followed by atherosclerotic infarctions (mean NIHSS score on admission 8). The mean NIHSS of ESUS patients was below the general NIHSS mean of 7. Lacunar infarctions showed even milder symptoms (mean NIHSS on admission 5) followed by TIA (mean NIHSS on admission 2), who had the lowest neurological deficits at discharge. From all stroke entities lacunar infarctions with a mean NIHSS score at discharge of 1 showed least neurological deficits. TIA patients were symptom-free after 24 hours, the NIHSS score here is 0. (Figure 2)

**Figure 2:**
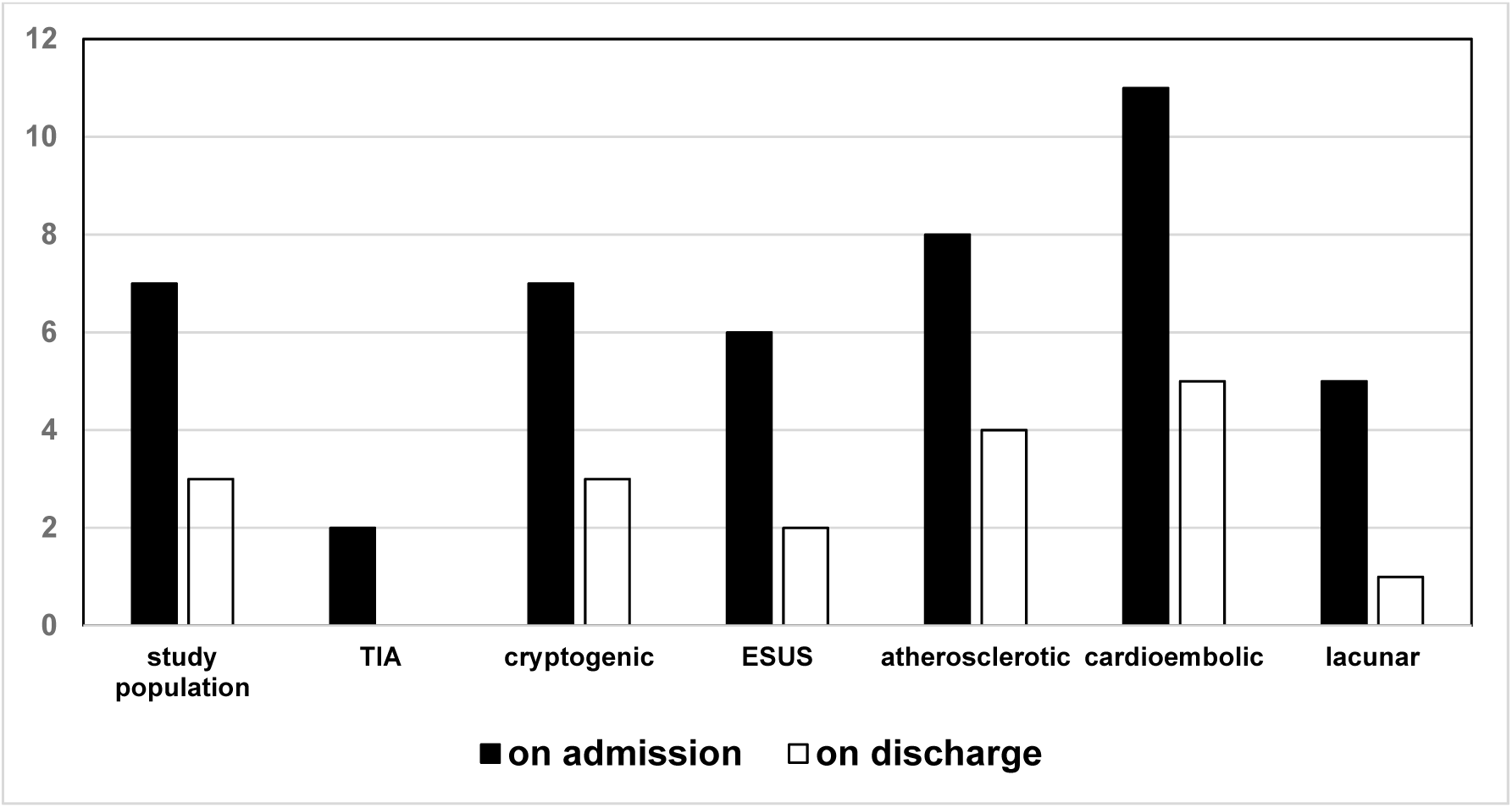
Mean NIHSS on admission and discharge in different stroke entities

### ECG monitoring and atrial fibrillation

By continuous heart rhythm monitoring we were able to detect any period of atrial fibrillation. According to current diagnostic criteria, atrial fibrillation is diagnosed with an episode lasting for more than 30 seconds. For a differentiated characterization, episodes of atrial fibrillation recorded under 30 seconds (i.e., 15-29 sec and 0-14 s) were analyzed and evaluated for different stroke entities.

Within the group of cardioembolic infarctions, 45% suffered from manifest atrial fibrillation (>30 s), 20% presented an atrial fibrillation episode of 15-29 s, and 22% an atrial fibrillation episode of 0-14 s. In total, more than 90% of cardioembolic infarcts showed an episode of atrial fibrillation of any duration. An AF episode of 15-29 s or 0-14 s was also found in almost 35% of the ESUS subgroup. This was similar in cryptogenic infarctions. 22% of TIA patients showed manifest atrial fibrillation, 7% showed one or more episodes of AF of 15-29 s and 10% an episode of 0-14 s. Among the atherosclerotic infarctions, the proportion of AF >30 s was lower at 12%, but the proportion of AF 15-29 sec was twice as high as shown in the TIA patients at 14%. The smallest proportion of atrial fibrillation of any duration is formed by patients with lacunar strokes, with a total of almost 25%, of which only 13% showed manifest AF.

In the overall population an episode of atrial fibrillation of any duration occurred in 54% of all stroke patients. Manifest atrial fibrillation shows a prevalence of 23% within the overall population, whereas 15% presented an AF episode of 15-29 s and 16% had an AF episode of 0-14 s.

**Figure 3:**
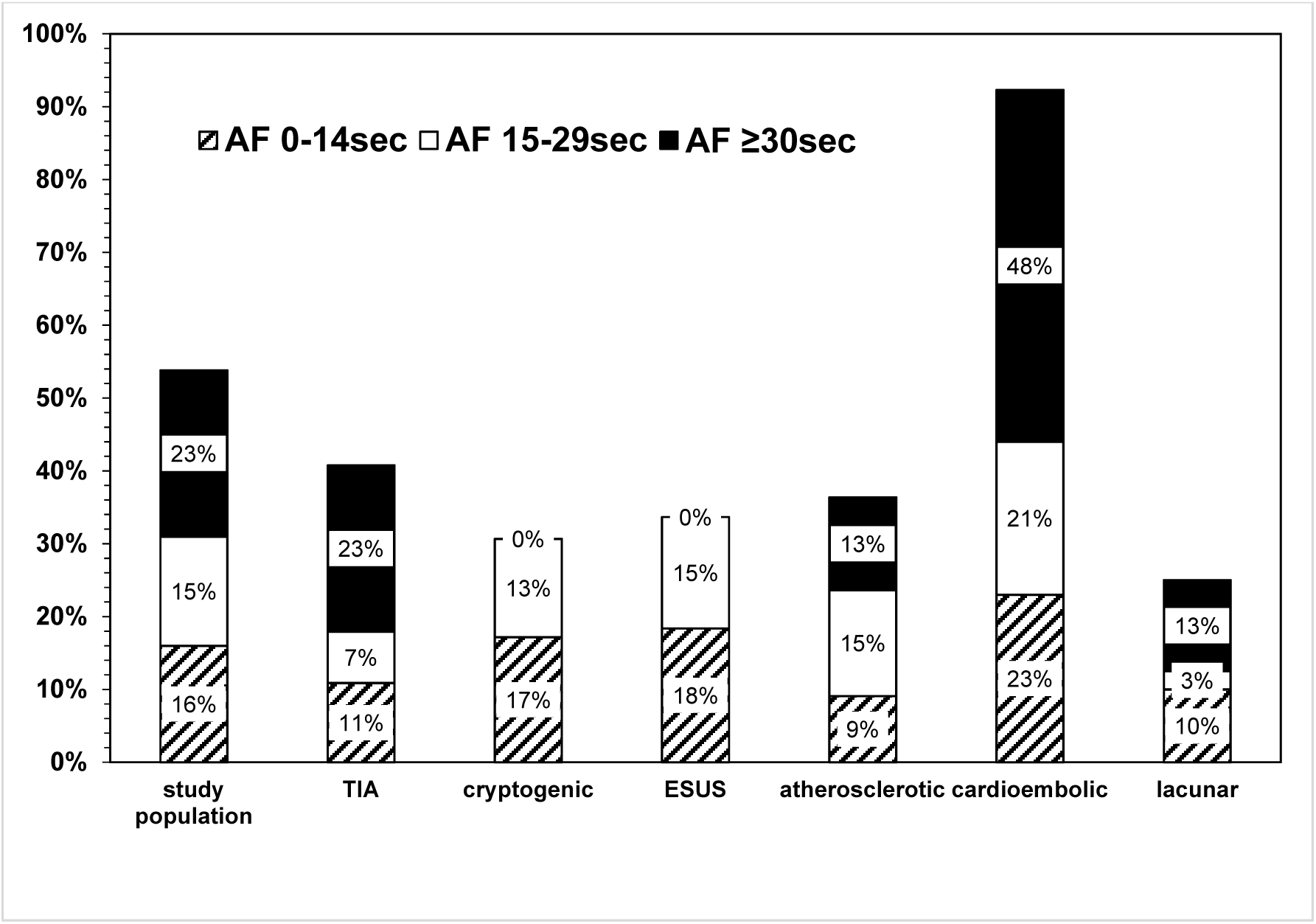
Episodes of AF > 30 and <30 seconds in the different stroke entities (+ TIA)

### CHA_2_DS_2_-VASC Score and short-term AF

To determine the risk of short-term AF patients we analyzed the common risk factor CHA_2_DS_2_-VASc score for each ischemic event group. Interestingly, among the all groups four fifth of the patients had a CHA_2_DS_2_-VASc score of 2-4 or higher (≥5). Among these groups 30-50% had an CHA_2_DS_2_-VASc score of 2-4, and 30-54% had a CHA_2_DS_2_-VASc score ≥5. However, lacunar stroke had 80% low risk patients, whereas all other groups had low risk patients within the rage of 15-22%.

**Figure 4:**
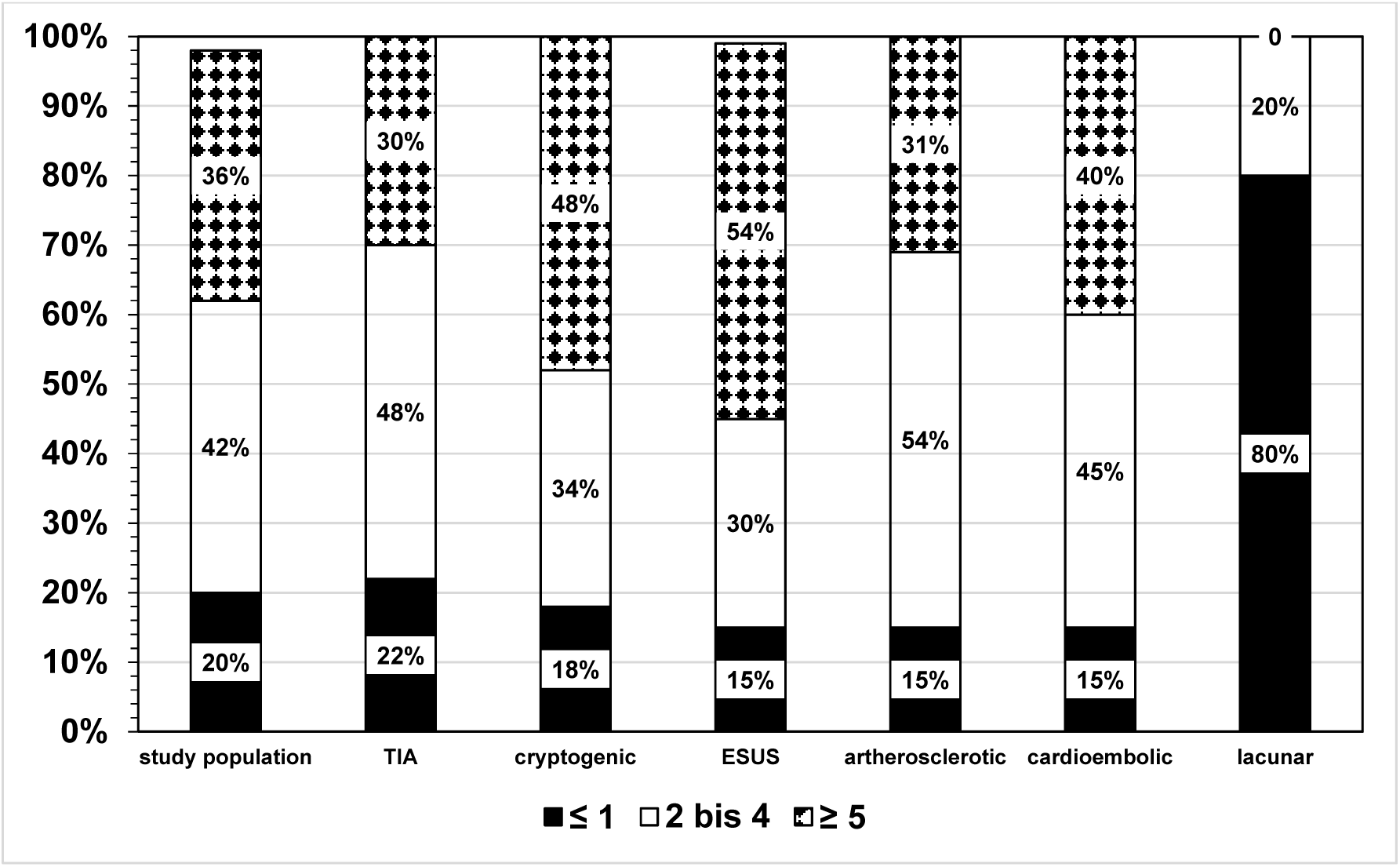
Analysis of CHA_2_DS_2_-VASc score in short term AF patients.

### Gender aspect of short-term AF

Analyzing the gender role in subclinical AF, we could detect short-term AF within a range of 25-71% in female patients suffering from ischemic cerebral events compared to a range of 10-44% in male patients. For the female study population compared to the male study population the difference was 37% vs. 23% and in total number 115 female patients vs. 92 male patients. Interestingly, female patients had twice the relative number of short-term AF compared to male patients.

**Figure 5.**
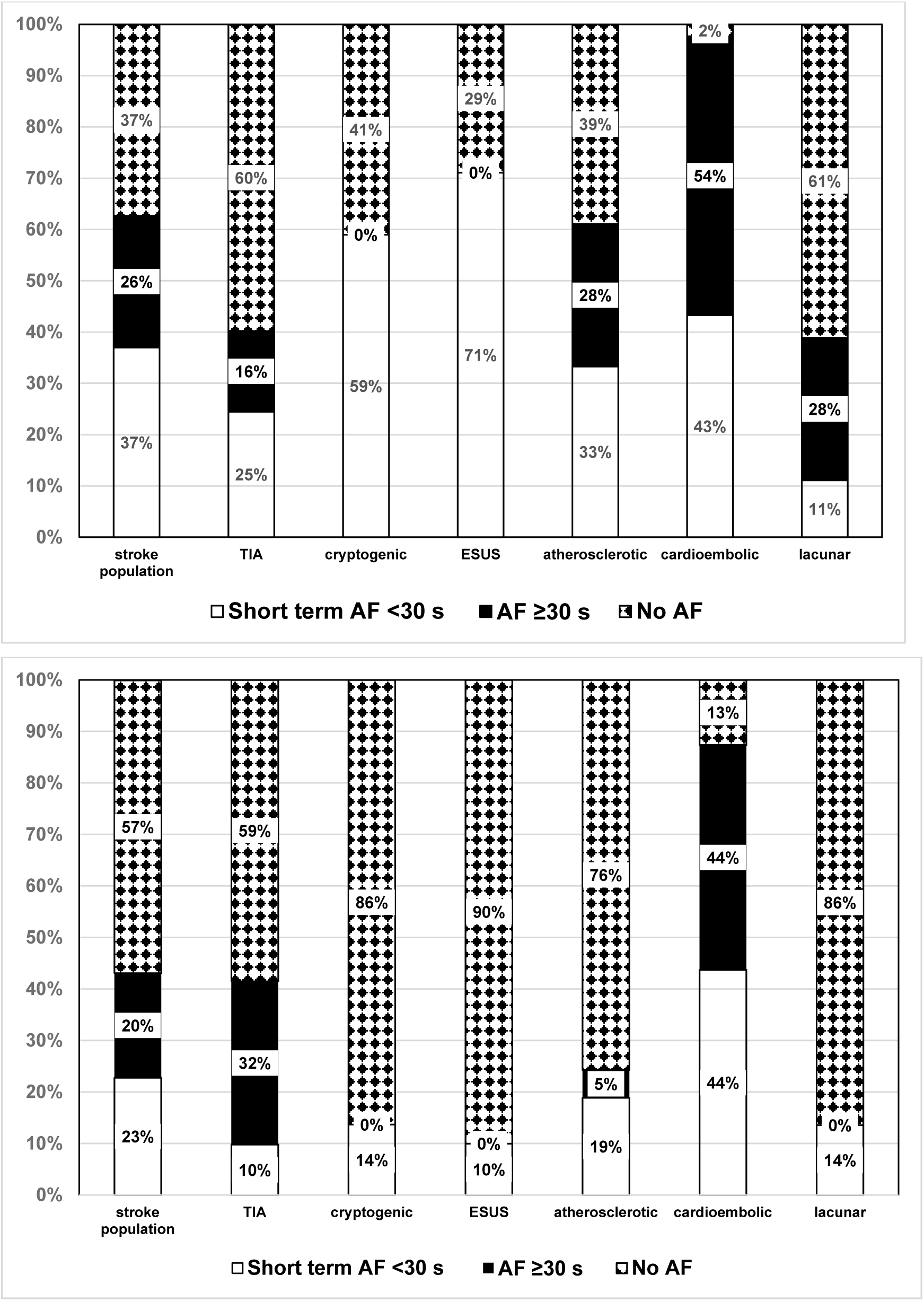
a+b: Analysis of short-term AF in female (a) and male (b) patients. Prevalence of short-term AF is twice as high in the female stroke population.

### Short term AF and age

The age analysis of patients with cerebral ischemic insults, showed that 60-79% of the patients with short-term AF were in the age groups 75-84 and ≥85 years. Whereas in the group of the age of 75-84 years there were twice as many patients with short-term AF than in the eldest age group. A higher age leads to a higher risk for short-term AF. This can be seen among all stroke entities. There was no major difference between the groups.

**Figure 6:**
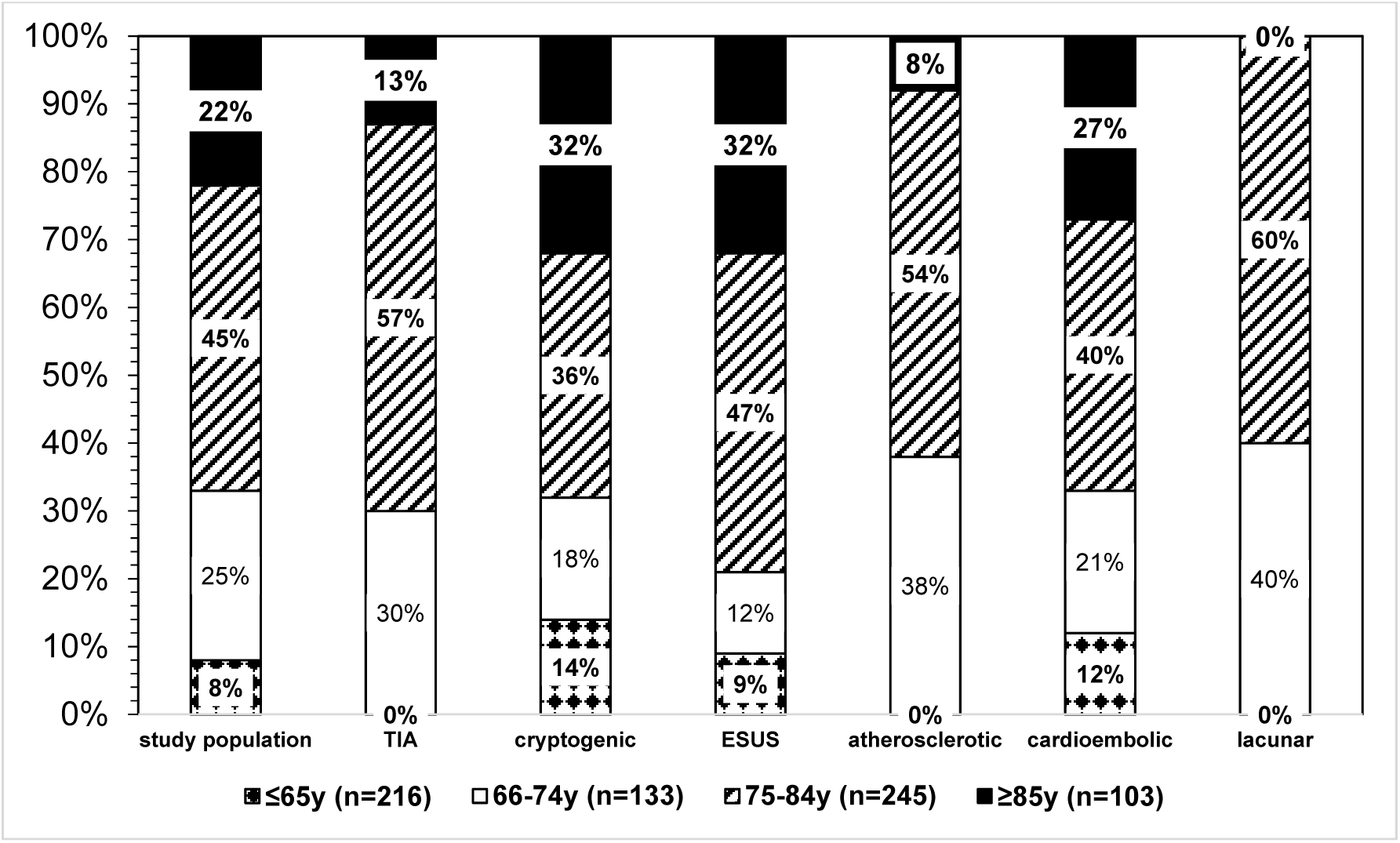
Analysis of age and short-term AF patients. Analysis of age in short-term AF patients showed that 2/3 of the patients were within the age groups of 75-84 and ≥85 years indicating the major role of age for AF development.

### Short term AF and severity of stroke

When we analyzed the severity of neurological injury with NIHSS in short term AF patients, we observed almost no symptoms (i.e., NIHSS 0) in the TIA group and cryptogenic/ESUS group. Severe neurological deficits (i.e., NIHSS 15-24, ≥25) were only seen in the cardioembolic group. So, the majority, 64-100% of the other stroke entities suffering from of the short-term AF had a mild to moderate neurologic injury (i.e., NIHSS= 1-5 or 6-14). This might indicate a fair recovery over time but also a profound threat of further damage due to recurrent stroke.

**Figure 7:**
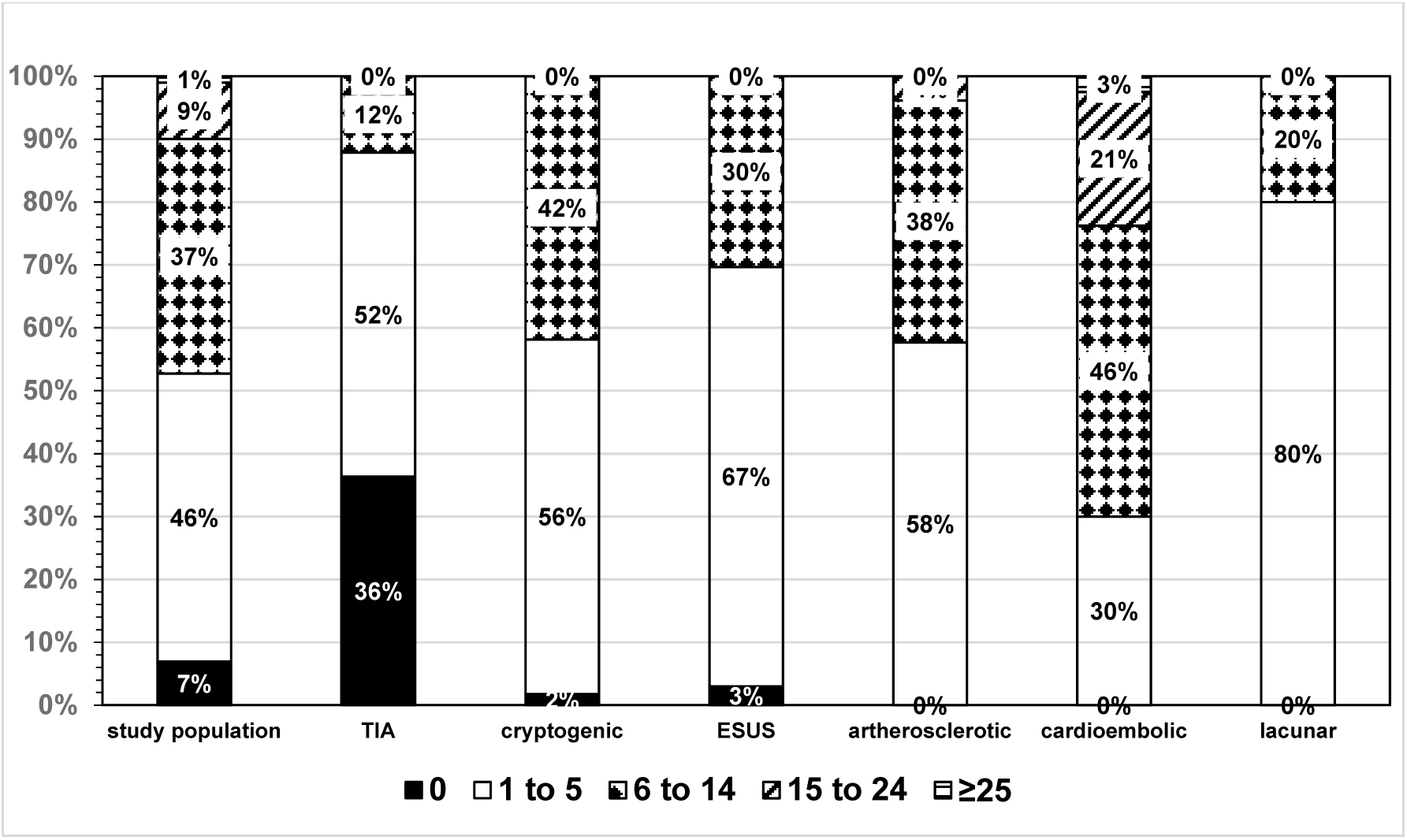
Analysis of short-term AF and severity of stroke (NIHHS) Analysis of short-term AF in patients with ischemic cerebral events shows 2/3 of the patients within in moderate neurologic injury severity (i.e., 1-5 and 6-14 NIHHS).

## DISCUSSION

With an incidence of 3.5% - 5.5%, ^20^ stroke is the third most frequent cause of death, with a probability of a second event of 12% per year. ^21^ In order to ensure optimal secondary prevention, the stroke etiology must be clarified very precisely. Despite the high medical standards in the industrialized nations, the etiology of 25-30% of all ischemic strokes remains unclear after completion of the inpatient diagnosis in the acute phase and is classified according to the TOAST criteria as called cryptogenic. ^10^ ^11^ Because these criteria did not set a minimum diagnostic standard to classify the stroke population, we additionally used the ESUS criteria from the "Cryptogenic Stroke/ESUS International Working Group” which categorized clear criteria and a minimum diagnostic standard for the etiological classification of a stroke. In this line, the identification of cardioembolic stroke patients is very important since therapeutic strategy differs in patients with or without AF. ECG, Holter ECG, ECG monitoring with automatic detection on strokes units and ICUs are used to identify AF in patients suffering from ischemic strokes.

In our study, all patients with an ischemic stroke or a TIA were prospectively included for 6 months and characterized using the ESUS criteria. The total population consists of 714 patients, of which 185 patients had a TIA. Among the stroke collective, 209 patients had a cardioembolic, 110 patients a atherosclerotic, 40 patients a lacunar stroke and 7 patients a stroke of other defined genesis. 163 patients had a cryptogenic stroke of which 98 met the minimum diagnostic standard for ESUS. In almost a quarter of the population, the stroke occurred as a second event.

## RISK FACTORS

Hypertension is the most prevalent risk factor in the stroke population with 75%, with >60% having insufficiently controlled hypertension. Nevertheless, with effective treatment a relative risk reduction of 30 to 40% can be archived. ^22^ The second most common risk factor was obesity at 50%, followed by hypercholesterolemia with 38%. Dominating the embolic risk factors, atrial fibrillation was most common in 48% of the cardioembolic strokes. In the ESUS subgroup, the most common embolic risk factor was CAD with 30%. Besides that, obesity had the highest relative frequency among ESUS patients at 55%.

## NEUROLOGICAL DEFICITS OF STROKE PATIENTS

All-cause mortality was 5%; among them >50% suffered from a cardioembolic infarction. In addition to the highest mortality, the subgroup also had the highest neurological deficits. 10% of cardioembolic infarctions showed severe to very severe neurological deficits (NIHSS >15) on admission and 28% still had moderate to very severe deficits at discharge. In comparison, 91% of ESUS patients showed mild to moderate symptoms on admission (NIHSS 0-14); of which 42% had no neurological deficits and 43% only had mild neurological deficits (NIHSS 0-5) at discharge. Only TIA patients and those with a lacunar infarct pattern showed little affection with the majority having no symptoms at discharge.

It should be noted that other studies recorded data retrospectively and the NIHSS score was based on doctor’s discharge letters or on the basis of recording sheets. In comparison, in our study, the NIHSS score was determined after presentation in the Emergency admission within a window of 0-24h by a study doctor based on the NIHSS sheet after a focused neurological examination.

### CHA_2_DS_2_ VASc-Score

When we divided the different stroke entities by the CHA_2_DS_2_-VASc (0-1, 2-5, ≥5) we noticed the largest proportion of a CHA_2_DS_2_-VASc score of ≥5 points in the cardioembolic group with approximately 40%. Previously it was shown that a high CHA_2_DS_2_-VASc might indicate future AF development. This coincides with the fact that over 50% of the patients with cardioembolic infarctions suffered from atrial fibrillation. The group with the second highest risk of suffering a recurrent stroke was the population having an atherosclerotic insult, of which 33% had a score >5 and 85% had a score of 2-4, followed by the ESUS and

cryptogenic stroke group, where just less than 30% had a score >5. Compared to the total population, the ESUS subgroup CHA_2_DS_2_-VASc score distribution was similar to the overall population. The distribution of the ESUS CHA_2_DS_2_-VASc score showed no relevant difference to other stroke entities.

### CASES OF COVERT ATRIAL FIBRILLATION – patients with short-term AF

Manifest atrial fibrillation (>30s) shows a prevalence within the overall population of 23%. In our study we have shown that 15% of the stroke patients had an AF episode of 15-29 s and 16% had an AF episode of 0-14 s. Nonvalvular atrial fibrillation in general is the most common cause of cardio cerebral embolism. ^23^ Equally, in our study, AF shows a frequency of 48% among cardioembolic infarctions. An AF episode of any duration exhibit nearly 90% in this group. Since manifest AF is an exclusion criterion for ESUS, none of the ESUS patients suffer from manifest AF. However, it is striking that 15% of ESUS patients had an AF episode of 15-29 s and 18% had an AF episode of 0-15 s. The short-term AF numbers are similar in the cryptogenic stroke subgroup. It remains unclear if these 33% of the ESUS group develop manifest AF in the future. However, when we analyze the CHA_2_DS_2_-VASc-Score, 80% of the short-term AF patients had a score of 2-4 or above, among them 30-50% having a score of ≥5 in the different stroke entities indicating a profound risk for a recurrent stroke. In the study from Saliba et al.^24^ it was shown that the CHA_2_DS_2_-VASc score corresponds with the risk for AF development increasing in patients having a score >2 points.

Similar when we looked on the gender aspect of short-term AF. In the male population short-term AF was prevalent in the different entities from 10-44%, whereas in the female group it was prevalent from 25-71%. Interestingly, stroke risk and therapeutic complications in women are much higher due to differences in risk factors, strength of risk factors, sex specific factors, yet they are less likely given therapy and underrepresented in clinical trials.^25^ Our results demonstrate the importance of short-term AF in women and possible future therapeutic aspects when earlier anticoagulation might be applied.

Further, when we determined the age aspect of short-term AF patients, we saw that the 2/3 of the patients with different strokes were in the range of 74-85 and ≥85 years of age. In addition, almost 95% of all stroke patients with short term AF are >65 years old. Clearly, age seems to play an important role, which is also represented in the risk stratification of the CHA_2_DS_2_-VASc score where patients receive 1 or 2 points for age. The association of AF with increasing age is well recognized and has been shown by several epidemiological studies. The risk of developing AF doubles with each progressive decade of gaining and exceeds 20% by age 80 years. Although important and well established, age is not the only risk factor associated with AF. ^26^ ^27^

When we analyzed the severity of strokes in short-term AF patients of different stroke entities, we recognized, that the majority of them experienced moderate stroke severity and recovered profoundly at discharge. However, when they were discharged with a platelet inhibitor there might be a high risk of recurrent strokes. In this line, Kim et al. ^28^ recently showed in a multivariable analysis that recurrence of any stroke was independently associated with AF diagnosed before the index stroke and lesion patterns of subcortical infarction and small scattered lesions in a single vascular territory. Similarly, Penado et al. ^29^ have shown that atrial fibrillation was an independent risk factor for stroke recurrence over a wide age range.

A source of cardioembolic infarction seems to exist in the ESUS collective and appears to have significant impact on future therapies. However, 30% of our cryptogenic stroke group and 33% of the ESUS group suffered from short-term AF. These patients had a moderate to high risk for developing a recurrent embolic stroke (i.e., CHA_2_DS_2_-VASc Score). However, in the RE-SPECT ESUS and NAVIGATE ESUS trial DOAC did not demonstrate any benefit. However, with the strategy of short-term AF we might be able to detect a subgroup among the ESUS patients which might benefit from OAC treatment.

Stroke severity was analyzed using the NIHSS. The majority of short-term AF patients (64-100%) had a moderate neurological injury (i.e. NIHSS of 1-5 or 6-14). This might indicate a fair recovery over time but also a profound threat of further damage due to recurrent stroke. Approximately 80% to 85% of the patients survive the first ischemic stroke, and among these, 15% to 30% experience a recurrent stroke within the first 2 years. ^11^ Recurrent stroke, as one of the clinical endpoints of the study mentioned above, is the main cause of death, re-hospitalization, and long-term disability. Compared to the first stroke, neurological impairment caused by recurrence is more severe, difficult to treat, and correlates with a higher mortality. Therefore, secondary prevention after the first stroke is of great significance to reduce recurrent strokes. A moderate NIHSS value during acute ischemic stroke correlates with the incidence of recurrent stroke. In the study by Zhou et al. 2020 ^30^ 10 risk factors were identified, that may aid clinicians in identifying high-risk patients for stroke recurrence. The important risk factors were diabetes mellitus, smoking status, peripheral artery disease, hypercoagulable state, depression, 24 h minimum systolic blood pressure, 24 h maximum diastolic blood pressure, age, family history of stroke, NIHSS score status.

Recently, Cameron et al. ^31^ have identified biomarkers for atrial fibrillation after stroke. This seems to be another option besides ECG monitoring. Comparably, Thijs et al. ^32^ have indicated that age and prolonged PR interval are associated with AF in cryptogenic stroke patients. In addition, Bahit et al. ^33^ revealed besides age, hypertension, higher body mass index, and diabetes, are independent predictors of AF after embolic stroke of undetermined source. In the study by Baturova et al. ^34^ high CHADS_2_ and CHA_2_DS_2_-VASc scores, predict new AF onset in Follow-up. Similarly, QRS duration might be considered a potential risk marker for prediction of AF after ischemic stroke. The SAFAS Study, Garnier et al. ^35^ suggests, that a multimodal approach combining imaging, electrocardiography and original biological markers resulted in good predictive models for AFDAS (AF Detected After Stroke). These results also suggest that AFDAS is probably related to an underlying atrial cardiopathy.

In conclusion, Sposato et al. ^36^ have claimed, that larger and sufficiently powered randomized controlled trials of prolonged cardiac monitoring assessing the risk of stroke recurrence are needed. And they call for further research on AFDAS and stroke recurrence, and a tailored approach when using prolonged cardiac monitoring after ischemic stroke or transient ischemic attack, focusing on patients at higher risk of AFDAS and, more importantly, at higher risk of cardiac embolism.

Understanding who is at higher risk of developing AF will assist in identifying patients who may benefit from more intense, long-term cardiac monitoring. In this line in our study, we have demonstrated that a large number of patients following stroke have short term AF (0-30 s) in combination with a significant number of risk factors (ie. Gender, AGE, and CHA_2_DS_2_-VASc Score) implying the profound risk for future AF and recurrent stroke. If this subgroup of all stroke entities might benefit from DOAC treatment to prevent future strokes have to be investigated in future studies.

## LIMITATIONS AND STRENGTHS

One methodological weakness of this work represents the monocentric orientation. Also existing oral anticoagulant or platelet aggregation inhibitor therapy would have been of interest. The outcome and follow-up of the stroke population would have been of interest after 12 and 24 months.

Strengths of our study are the prospective work and standardized recording. In the majority of stroke and ESUS studies data was collected retrospectively and might be incomplete. Our stroke patient monitoring and clinical evaluation was done daily and done prospectively while the patient was hospitalized in a standardized clinical examination manner. Furthermore, precise ECG monitoring and analysis was performed daily.

## SUMMARY AND CONCLUSION

In summary, short-term AF is predominant in all stroke entities, being most prevalent in cardioembolic strokes. In more than 30% of the ESUS and cryptogenic stroke patients short-term AF could be detected during hospitalization. Possibly, the definition of AF as defined by the ESC has to be adjusted in the future for patients with a high cardioembolic risk. All short-term AF subgroups had a CHA_2_DS_2_-VASc Score of a minimum of 2 points, and a moderate stroke severity (NIHSS 1-14) and might benefit from DOAC treatment to prevent recurrent strokes. Elderly patients and women seem to be at higher risk for presenting short-term AF and developing manifest AF in the future.

## Data Availability

data on file

## LITERATURE

1. Flaker GC, Belew K, Beckman K, Vidaillet H, Kron J, Safford R, Mickel M, Barrell P, Investigators A. Asymptomatic atrial fibrillation: demographic features and prognostic information from the Atrial Fibrillation Follow-up Investigation of Rhythm Management (AFFIRM) study. Am Heart J. 2005;149:657–663. doi: 10.1016/j.ahj.2004.06.032

2. Dilaveris PE, Kennedy HL. Silent atrial fibrillation: epidemiology, diagnosis, and clinical impact. Clin Cardiol. 2017;40:413–418. doi: 10.1002/clc.22667

3. Wolf PA, Dawber TR, Thomas HE, Jr., Kannel WB. Epidemiologic assessment of chronic atrial fibrillation and risk of stroke: the Framingham study. Neurology. 1978;28:973–977. doi: 10.1212/wnl.28.10.973

4. Wolf PA, Abbott RD, Kannel WB. Atrial fibrillation: a major contributor to stroke in the elderly. The Framingham Study. Arch Intern Med. 1987;147:1561–1564.

5. Liao J, Khalid Z, Scallan C, Morillo C, O’Donnell M. Noninvasive cardiac monitoring for detecting paroxysmal atrial fibrillation or flutter after acute ischemic stroke: a systematic review. Stroke. 2007;38:2935–2940. doi: 10.1161/STROKEAHA.106.478685

6. Calkins H, Hindricks G, Cappato R, Kim YH, Saad EB, Aguinaga L, Akar JG, Badhwar V, Brugada J, Camm J, et al. 2017 HRS/EHRA/ECAS/APHRS/SOLAECE expert consensus statement on catheter and surgical ablation of atrial fibrillation: Executive summary. Heart Rhythm. 2017;14:e445–e494. doi: 10.1016/j.hrthm.2017.07.009

7. Charitos EI, Stierle U, Ziegler PD, Baldewig M, Robinson DR, Sievers HH, Hanke T. A comprehensive evaluation of rhythm monitoring strategies for the detection of atrial fibrillation recurrence: insights from 647 continuously monitored patients and implications for monitoring after therapeutic interventions. Circulation. 2012;126:806–814. doi: 10.1161/CIRCULATIONAHA.112.098079

8. Adams HP, Jr., Bendixen BH, Kappelle LJ, Biller J, Love BB, Gordon DL, Marsh EE, 3rd. Classification of subtype of acute ischemic stroke. Definitions for use in a multicenter clinical trial. TOAST. Trial of Org 10172 in Acute Stroke Treatment. Stroke. 1993;24:35–41. doi: 10.1161/01.str.24.1.35

9. Pinto A, Tuttolomondo A, Di Raimondo D, Fernandez P, Licata G. Risk factors profile and clinical outcome of ischemic stroke patients admitted in a Department of Internal Medicine and classified by TOAST classification. Int Angiol. 2006;25:261–267.

10. Hart RG, Diener HC, Coutts SB, Easton JD, Granger CB, O’Donnell MJ, Sacco RL, Connolly SJ, Cryptogenic Stroke EIWG. Embolic strokes of undetermined source: the case for a new clinical construct. Lancet Neurol. 2014;13:429–438. doi: 10.1016/S1474-4422(13)70310-7

11. Grau AJ, Weimar C, Buggle F, Heinrich A, Goertler M, Neumaier S, Glahn J, Brandt T, Hacke W, Diener HC. Risk factors, outcome, and treatment in subtypes of ischemic stroke: the German stroke data bank. Stroke. 2001;32:2559–2566. doi: 10.1161/hs1101.098524

12. Ntaios G, Papavasileiou V, Milionis H, Makaritsis K, Vemmou A, Koroboki E, Manios E, Spengos K, Michel P, Vemmos K. Embolic Strokes of Undetermined Source in the Athens Stroke Registry: An Outcome Analysis. Stroke. 2015;46:2087–2093. doi: 10.1161/STROKEAHA.115.009334

13. Li F, Yang L, Yang R, Xu W, Chen FP, Li N, Zhang JB. Ischemic Stroke in Young Adults of Northern China: Characteristics and Risk Factors for Recurrence. Eur Neurol. 2017;77:115–122. doi: 10.1159/000455093

14. Hindricks G, Potpara T, Dagres N, Arbelo E, Bax JJ, Blomstrom-Lundqvist C, Boriani G, Castella M, Dan GA, Dilaveris PE, et al. 2020 ESC Guidelines for the diagnosis and management of atrial fibrillation developed in collaboration with the European Association for Cardio-Thoracic Surgery (EACTS): The Task Force for the diagnosis and management of atrial fibrillation of the European Society of Cardiology (ESC) Developed with the special contribution of the European Heart Rhythm Association (EHRA) of the ESC. Eur Heart J. 2021;42:373–498. doi: 10.1093/eurheartj/ehaa612

15. Diener HC, Sacco RL, Easton JD, Granger CB, Bernstein RA, Uchiyama S, Kreuzer J, Cronin L, Cotton D, Grauer C, et al. Dabigatran for Prevention of Stroke after Embolic Stroke of Undetermined Source. N Engl J Med. 2019;380:1906–1917. doi: 10.1056/NEJMoa1813959

16. Hart RG, Sharma M, Mundl H, Kasner SE, Bangdiwala SI, Berkowitz SD, Swaminathan B, Lavados P, Wang Y, Wang Y, et al. Rivaroxaban for Stroke Prevention after Embolic Stroke of Undetermined Source. N Engl J Med. 2018;378:2191–2201. doi: 10.1056/NEJMoa1802686

17. Brott T, Adams HP, Jr., Olinger CP, Marler JR, Barsan WG, Biller J, Spilker J, Holleran R, Eberle R, Hertzberg V, et al. Measurements of acute cerebral infarction: a clinical examination scale. Stroke. 1989;20:864–870. doi: 10.1161/01.str.20.7.864

18. Goldstein LB, Samsa GP. Reliability of the National Institutes of Health Stroke Scale. Extension to non-neurologists in the context of a clinical trial. Stroke. 1997;28:307–310. doi: 10.1161/01.str.28.2.307

19. Meyer BC, Lyden PD. The modified National Institutes of Health Stroke Scale: its time has come. Int J Stroke. 2009;4:267–273. doi: 10.1111/j.1747-4949.2009.00294.x

20. Feigin VL, Lawes CM, Bennett DA, Anderson CS. Stroke epidemiology: a review of population-based studies of incidence, prevalence, and case-fatality in the late 20th century. Lancet Neurol. 2003;2:43–53. doi: 10.1016/s1474-4422(03)00266-7

21. Petty GW, Brown RD, Jr., Whisnant JP, Sicks JD, O’Fallon WM, Wiebers DO. Survival and recurrence after first cerebral infarction: a population-based study in Rochester, Minnesota, 1975 through 1989. Neurology. 1998;50:208–216. doi: 10.1212/wnl.50.1.20822.

22. Hong KS. Blood Pressure Management for Stroke Prevention and in Acute Stroke. J Stroke. 2017;19:152–165. doi: 10.5853/jos.2017.00164

23. Arboix A, Alio J. Cardioembolic stroke: clinical features, specific cardiac disorders and prognosis. Curr Cardiol Rev. 2010;6:150–161. doi: 10.2174/157340310791658730

24. Saliba W, Gronich N, Barnett-Griness O, Rennert G. Usefulness of CHADS2 and CHA2DS2-VASc Scores in the Prediction of New-Onset Atrial Fibrillation: A Population-Based Study. Am J Med. 2016;129:843–849. doi: 10.1016/j.amjmed.2016.02.029

25. Rexrode KM, Madsen TE, Yu AYX, Carcel C, Lichtman JH, Miller EC. The Impact of Sex and Gender on Stroke. Circ Res. 2022;130:512–528. doi: 10.1161/CIRCRESAHA.121.319915

26. Lernfelt G, Mandalenakis Z, Hornestam B, Lernfelt B, Rosengren A, Sundh V, Hansson PO. Atrial fibrillation in the elderly general population: a 30-year follow-up from 70 to 100 years of age. Scand Cardiovasc J. 2020;54:232–238. doi: 10.1080/14017431.2020.1729399

27. Kornej J, Borschel CS, Benjamin EJ, Schnabel RB. Epidemiology of Atrial Fibrillation in the 21st Century: Novel Methods and New Insights. Circ Res. 2020;127:4–20. doi: 10.1161/CIRCRESAHA.120.316340

28. Kim BJ, Hwang YH, Park MS, Kim JT, Choi KH, Jung JM, Yu S, Kim CK, Oh K, Song TJ, et al. Atrial Fibrillation Related and Unrelated Stroke Recurrence Among Ischemic Stroke Patients With Atrial Fibrillation. Front Neurol. 2021;12:744607. doi: 10.3389/fneur.2021.744607

29. Penado S, Cano M, Acha O, Hernandez JL, Riancho JA. Atrial fibrillation as a risk factor for stroke recurrence. Am J Med. 2003;114:206–210. doi: 10.1016/s0002-9343(02)01479-1

30. Zhuo Y, Wu J, Qu Y, Yu H, Huang X, Zee B, Lee J, Yang Z. Clinical risk factors associated with recurrence of ischemic stroke within two years: A cohort study. Medicine (Baltimore). 2020;99:e20830. doi: 10.1097/MD.0000000000020830

31. Cameron A, Cheng HK, Lee RP, Doherty D, Hall M, Khashayar P, Lip GYH, Quinn T, Abdul-Rahim A, Dawson J. Biomarkers for Atrial Fibrillation Detection After Stroke: Systematic Review and Meta-analysis. Neurology. 2021;97:e1775–e1789. doi: 10.1212/WNL.0000000000012769

32. Thijs VN, Brachmann J, Morillo CA, Passman RS, Sanna T, Bernstein RA, Diener HC, Di Lazzaro V, Rymer MM, Hogge L, et al. Predictors for atrial fibrillation detection after cryptogenic stroke: Results from CRYSTAL AF. Neurology. 2016;86:261–269. doi: 10.1212/WNL.0000000000002282

33. Bahit MC, Sacco RL, Easton JD, Meyerhoff J, Cronin L, Kleine E, Grauer C, Brueckmann M, Diener HC, Lopes RD, et al. Predictors of Atrial Fibrillation Development in Patients With Embolic Stroke of Undetermined Source: An Analysis of the RE-SPECT ESUS Trial. Circulation. 2021;144:1738–1746. doi: 10.1161/CIRCULATIONAHA.121.055176

34. Baturova MA, Lindgren A, Carlson J, Shubik YV, Olsson SB, Platonov PG. Predictors of new onset atrial fibrillation during 10-year follow-up after first-ever ischemic stroke. Int J Cardiol. 2015;199:248–252. doi: 10.1016/j.ijcard.2015.07.047

35. Garnier L, Duloquin G, Meloux A, Benali K, Sagnard A, Graber M, Dogon G, Didier R, Pommier T, Vergely C, et al. Multimodal Approach for the Prediction of Atrial Fibrillation Detected After Stroke: SAFAS Study. Front Cardiovasc Med. 2022;9:949213. doi: 10.3389/fcvm.2022.949213

36. Sposato LA, Chaturvedi S, Hsieh CY, Morillo CA, Kamel H. Atrial Fibrillation Detected After Stroke and Transient Ischemic Attack: A Novel Clinical Concept Challenging Current Views. Stroke. 2022;53:e94–e103. doi: 10.1161/STROKEAHA.121.034777

